# A radiomics approach to artificial intelligence in echocardiography: Predicting post-operative right ventricular failure

**DOI:** 10.1101/2020.05.05.20092494

**Authors:** Rohan Shad, Nicolas Quach, Robyn Fong, Curt P. Langlotz, Sandra Kong, Patpilai Kasinpila, Myriam Amsallem, Francois Haddad, Yasuhiro Shudo, Y. Joseph Woo, Jeffrey Teuteberg, William Hiesinger

## Abstract

In this study, we describe a novel ‘radiomics’ approach to an echocardiography artificial intelligence system that enables the extraction of hundreds of thousands of motion parameters per echocardiography video. We apply this AI system to the clinical problem of predicting post-operative right ventricular failure (RV failure) in heart failure patients receiving implantable circulatory life support systems. Post-operative RV failure is the single largest contributor to short-term mortality in patients with left ventricular assist devices (LVAD); yet predicting which patient is at risk of developing this complication in the pre-operative setting, has remained beyond the abilities of experts in the field. We report results on testing datasets using a standard 10-fold cross validation. The AUC for the AI system trained using the Stanford LVAD dataset was 0.860 (95% CI 0.815-0.905; n = 290 patients) using pre-operative echocardiograms alone. We further show that our system outperforms board certified clinicians equipped with both contemporary risk scores (AUC 0.502 - 0.584) and independently measured echocardiographic metrics (0.519 – 0.598).

## Introduction

In recent years, artificial intelligence has enabled automated systems to meet or exceed the performance of clinical experts across a range of image analysis tasks.^1-4^ Echocardiography video analysis poses both new challenges and opportunities – while the presence of a temporal dimension increases the complexity of the problem, the density of information in videos enables translation of artificial intelligence techniques to patient populations with comparatively limited datasets.^3^ All current automated echocardiography systems – much like human echocardiography reads – are inherently reductionist in nature; a complex sequence and pattern of cardiac contraction is reduced to an outline of one or more chambers, from which a few global metrics of heart function are then calculated.^5,6^ Extracting the subtle motion characteristics of the heart that may be predictive of diseased states thus requires a fundamental shift in approach to AI in echocardiography.

Heart failure affects more than 6.5 million people in the United States alone, with an estimated 960,000 new cases diagnosed each year.^7^ A heart transplant remains the gold standard for treating patients with end-stage heart failure. Demand, however, far outpaces the supply of transplantable hearts.^8,9^ Left ventricular assist devices (LVADs) offer a mechanical alternative to transplantation, and the number of patients supported by these battery powered mechanical pumps have steadily grown since 2008.^10^ In the contemporary era, an estimated 3500 LVAD implants are performed each year, with 10 year outcomes comparable to transplants.^11,12^ Unfortunately, approximately a third of all patients implanted with LVADs, develop a clinically significant degree of right ventricular failure (RV failure) soon after the procedure.^13,14^ Severe RV failure remains the single largest contributor to short-term mortality in this patient population.^11,15,16^

Predicting which patient will go on to develop RV failure has so far remained beyond the abilities of experts in the field. A variety of clinical scoring systems have been developed with modest predictive power, with an area under curve (AUC) of the receiver operating (ROC) curve of 0.65 at best (Figure 1).^13,17-20^ Without reliable methods to predict RV failure in the pre-operative setting, we have neither the means to decide in whom to aggressively intervene, nor a way to randomize patients to trials that evaluate the efficacy of right ventricular treatment options. The gold standard for determining which patients should receive advanced right ventricular support devices thus remains a ‘clinical gestalt’,^21^ involving the patients’ clinical course, lab parameters, and a qualitative assessment of myocardial function using a transthoracic echocardiogram.

In this study, we describe a novel ‘radiomics’ approach to an echocardiography artificial intelligence system that enables the extraction of hundreds of thousands of motion parameters per ECHO. We then use this AI system for the prediction of post-operative RV failure in LVAD patients, using pre-operative ECHOs alone. We compare the predictions of our AI system to those of contemporary RV failure risk scores and further show that the performance of our system transcends human capabilities. Finally, we use a series of strategies to interpret and visualize how our algorithms function, drawing from them pathophysiological insights into RV failure in LVAD patients. Figure 1 details an overview of the project.

## Methods

### Data sources and study population

Data in the form of clinical outcomes and raw echocardiography DICOM files were sourced from the department of Cardiothoracic Surgery, Stanford University (IRB 52440). All patients aged 18-years or older with at least one pre-operative transthoracic echocardiogram as well as a complete pre-operative and post-operative assessment of RV failure during index-hospitalization as per the INTERMACS adverse events definitions (Interagency Registry for Mechanically Assisted Circulatory Support) were included (Supplementary Table 1).^22^ Apical 4 chamber views for trans-thoracic echocardiograms taken closest to the day of surgery were used for this study. Our total dataset comprised 290 patients. Raw data was anonymized and linked to clinical outcomes data via a unique study-ID. Post-operative RV failure was defined by INTERMACS defined criteria. As these definitions were standardized later in 2014, we manually reviewed each patient record for the duration of inpatient admission to collect pre-operative and post-operative clinical parameters following a pre-determined and standardized protocol. This enabled grading of RV failure severity along with the calculations of the various RV failure risk scores. Clinical data was stored and managed in REDCap.

### Outcomes

The primary outcome of the study was the ability of the machine learning pipeline to identify and predict the likelihood of post-operative RV failure using only pre-operative transthoracic echocardiograms as the input. All patients with severe or higher grades of post-operative RV failure were included in the ‘RV failure’ group (n = 106 patients; 36%); the remainder were kept as controls (n = 184). Multiple ECHOs were available for each patient (297 RV failure (34%); 569 normal). The performance was mathematically assessed by the area under curve (AUC) of the receiver operating characteristic (ROC) curve, precision-recall curves, and balanced accuracy of the algorithm.

**Fig. 1.**
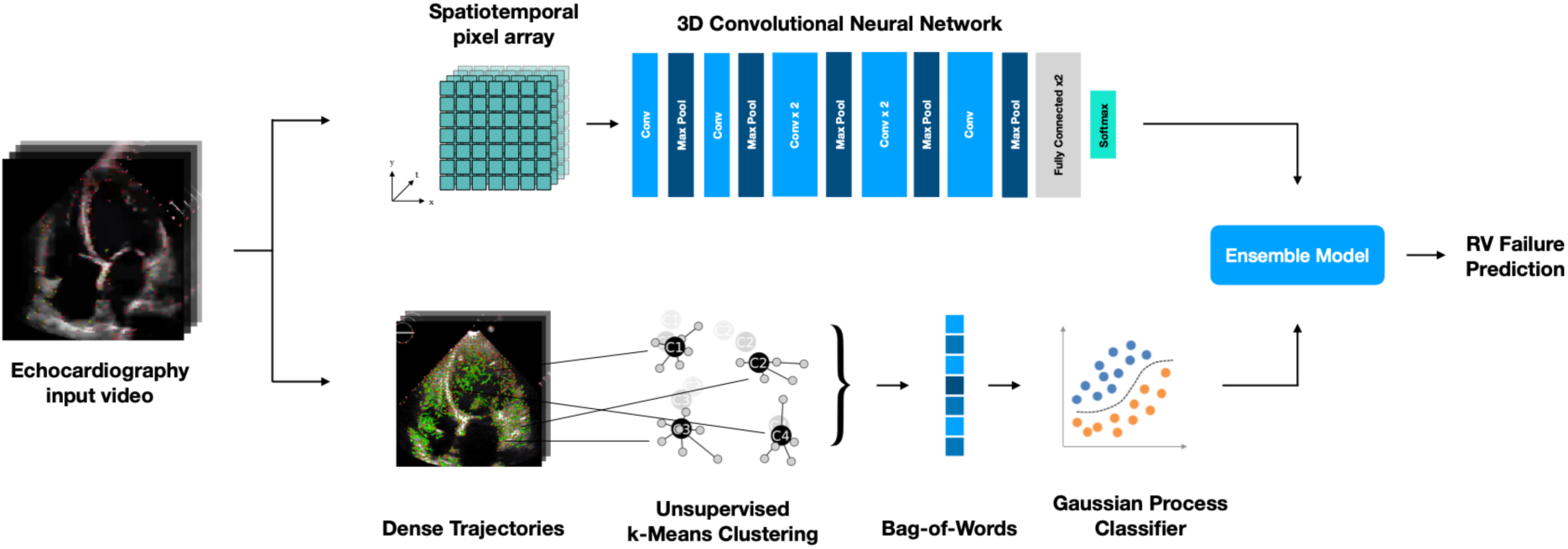
Schematic of AI echocardiography pipeline. The input is an apical 4 chamber transthoracic echocardiogram, which is processed by two parallel algorithms: A supervised 3D convolutional neural network, and an unsupervised improved dense trajectory system paired with a supervised classifier for predicting the outcome of RV failure. The final prediction of RV failure is made by combining the predictions from each half of the pipeline with an ensemble model.

### Data pre-processing

Echocardiograms were first de-identified by stripping all private health information (PHI) from file metadata and by obscuring any sensitive information in the videos. The complete removal of all sensitive information was verified manually on all videos before proceeding to downstream postprocessing. Areas outside of the scanning sector were masked to remove any miscellaneous markings in the video frames that may otherwise influence the neural networks. The videos were then normalized by dividing each pixel value by the pixel of maximal intensity. The processed videos at their original resolution were used for the improved dense trajectory pipeline. For the 3D conv-net pipeline, the frames of the processed videos were additionally down-sampled by bi-linear interpolation to a 112×112 resolution for training.

### Machine learning pipeline

Our automated echocardiography analysis pipeline is a combination of two algorithms. The first algorithm is an improved dense trajectory system, originally developed for action recognition in large scale video datasets, that tracks motion features across short time intervals.^23^ The second is a 3-dimensional convolutional neural network,^24^ built using the Keras Framework with a TensorFlow (Google; Mountain View, CA, USA) backend and Python, that tracks motion features and structural features over multiple cardiac cycles. Improved dense trajectories were first described by Wang *et al* to densely sample thousands of feature points from each video, tracking them across time using optical flow. Six ‘trajectory descriptors’ representing information of shape, appearance, and motion are calculated for each tracked feature point. The totality of local spatiotemporal information is reduced to a bag-of-words representation using an unsupervised k-means clustering algorithm, following which a probabilistic prediction of RV failure outcome is produced by a supervised Gaussian Process Classifier.

The 3-dimensional convolutional neural network consists of 8 convolutional layers, 5 max-pooling layers, and 2-fully connected layers that culminate into a final softmax classifier for RV failure prediction (Fig 1.). All convolutional layers utilize a 3×3×3 kernel, except for the first convolutional layer which does not convolve over the temporal dimension. The network weights were initialized using the Xavier normal initializing scheme, and was optimized using the Adam algorithm.^25,26^ The network was trained for 25 epochs on a batch size of 8, with an initial learning rate of 1×10^-5^. Training was stopped early if the loss did not improve for 5 epochs. For each echocardiogram, 5 random 64-frame clips of the full movie were subsampled and passed through the trained neural network. The average of the 5 outputs was calculated to predict RV failure. The predicted probabilities of RV failure from both the dense trajectory analysis and convolutional neural network were ensembled using a weighted soft-voting classifier. Here the probabilities for each outcome class is calculated for the two halves of the AI system, and an empirically derived weight is applied to each probability and then summed together. The outcome class with the highest weighted sum is outputted as the predicted label. The models were trained on servers, each with eight NVIDIA V100 GPUs, on the Stanford Sherlock Supercomputing Cluster.

### Visualizations and interpretations

We used an implementation of Layerwise relevance propagation to highlight regions of each ECHO being used to derive predictions of RV failure outcome when passed through the 3-dimensional convolutional neural network.^27^ We additionally visualized the bag-of-words representations of improved dense trajectory descriptors using pairwise correlation heatmaps, for patients with and without RV failure.

### Statistical analyses

To evaluate the stand-alone performance of the AI system, ROC curves were calculated as empirical curves in the sensitivity and specificity space. AUCs for ROC curves were computed with trapezoids using the pROC package.^28^ AUC for the Precision-Recall curve was computed using the interpolation method described by Davis and Goadrich.^29^ To compare the performance of our AI system against clinical risk scores and manually calculated echo metrics, we calculated non-parametric confidence intervals on the AUC using DeLong’s method,^30^ following which p-values were computed for the mean difference between AUC curves. Statistical analyses were conducted in R (v3.6.2).

### AI performance and comparison with clinical risk scores

In order to evaluate the performance of our echocardiogram analysis pipeline, a standard 10-fold cross validation was performed. The dataset was split in a 90:10 ratio into a training and test set, following which the pipeline was trained on the training set. The trained model was validated on the test set. This was repeated for a total of 10 trials, each time using a unique 90:10 split of the full dataset. The average performance on the test-datasets from each of these 10 trials was taken as the overall performance. The performance of the model was measured using balanced accuracy, area under curve (AUC) of the receiver-operator characteristic, AUC of the precision-recall curve, and F1 score. We further compared the predictive performance of our AI system against two contemporary risk scores used in for predicting post-operative RV failure - the CRITT score and Penn score. Missing data prevented the calculation of scores for 61 patients. As a result, further analyses were conducted on the remaining 229 patients. The variables used for calculating these scores are described in Supplementary Table 2.^13,18^

**Fig. 2:**
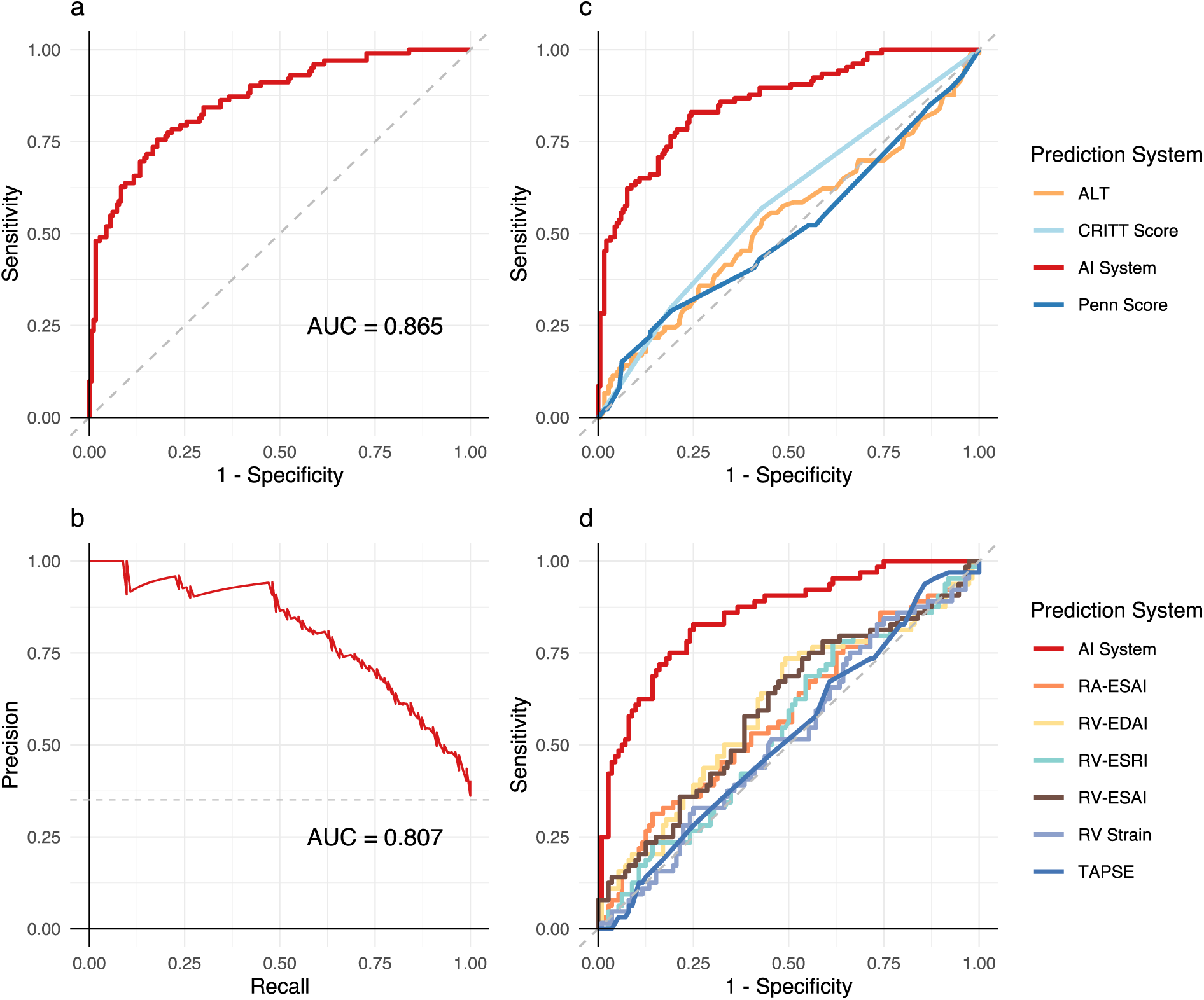
Performance of the AI echocardiography system, clinical risk scores, and manually calculated right heart functional metrics. **a**. Receiver operating characteristic curve for the AI system; and **b**. Precision-Recall curve for the AI system (n = 290). **c**. The ROC curves of contemporary state of the art clinical risk scores (n = 229), and **d**. The ROC curve for the AI system compared to an independently calculated set of echo metrics for right heart function (n = 176).

### Comparisons with manually calculated echocardiography metrics

We compared the performance of AI based RV failure prediction to a set of manually derived echocardiographic measures of right ventricular function. These measures were independently calculated by two board certified cardiologists for 176 patients (n = 112 controls; n = 64 with RV failure) in our dataset, using metrics of RV function that they have described in the past.^31^

## Results

The ROC and Precision-Recall curve of the AI system on the Stanford LVAD dataset are shown in Fig 2a and b, The ROC AUC for the AI system was 0.860 (95% CI 0.815-0.905; n = 290 patients). The AUCs calculated for the Penn score (0.502; 95% CI 0.423-0.582) and the CRITT score (0.584; 95% CI 0.509-0.653) for our dataset are similar to previously published reports (Fig 2c).^13,20^ Our AI system exceeded the performance of the best performing clinical risk score by a significant margin (change in area under curve (∆AUC) = +0.278, 95% CI 0.262-0.304, p < 0.0001, n = 229).

The AI system exceeded the performance of an independently calculated list of echocardiographic metrics of right ventricular load dependency. The AUCs of the manually extracted metrics ranged between 0.519 – 0.598. The AUC of the AI system on this subset of 176 patients was 0.850 (95% CI 0.791-0.909), compared to the best performing manual echo metric with an AUC of 0.598 (95% CI 0.509-0.688; ∆AUC 0.251 (95% CI 0.221-0.282, p < 0.0001) (Fig 2d).

Layerwise relevance propagation visualizations showed that for each patient, regions of activation for the 3d convolutional neural network were localized exclusively to the myocardium and valves. The cardiac chambers themselves showed no activation. Furthermore, motion characteristics of different regions of the heart contribute towards the prediction of RV failure at different phases of the cardiac cycle (Fig 3).

## Discussion

In this study we present a novel AI echocardiography system capable of extracting subtle myocardial motion aberrations for downstream clinical analyses. We utilize this system to predict the outcome of post-operative RV failure in 290 heart failure patients considered for LVAD implant, and further show that our AI echocardiography system outperforms board certified clinicians equipped with both manually extracted echo metrics and state of the art clinical risk scores. In the current body of work, we predict a binary outcome of RV failure, though our methods can readily be extended to predict either continuous or multi-class outcomes of interest.

**Fig. 3:**
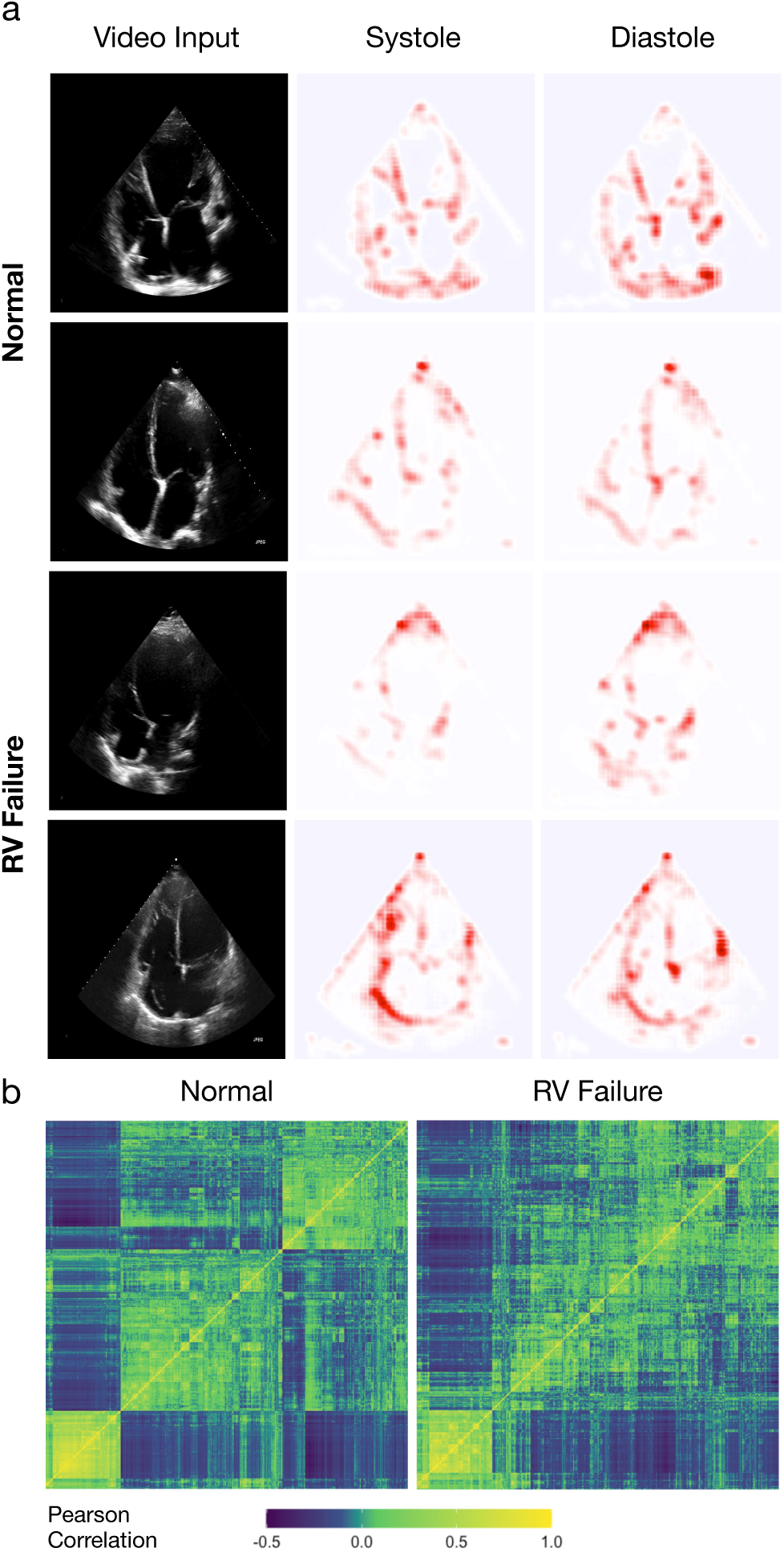
Radiomics approach to echocardiography AI visualized. **a**. Representative input ECHOs for the 3d-convoutional neural network, and Layerwise relevance propagation visualizations for both systolic and diastolic phases of the cardiac cycle across patients with and without RV failure. **b**. Heatmaps generated for the ‘trajectory’ descriptor from the improved dense trajectory pipeline, visualizing the subtle motion features tracked in normal vs RV failure cases.

All contemporary echocardiography AI systems rely on various supervised segmentation algorithms to outline cardiac chambers. Most recently, Ouyang *et al* describe an automated segmentation based system to calculate ejection fraction using left ventricular tracings.^6^ Our methods offer a number of key improvements: First, instead of cardiac chambers, our AI system extracts spatiotemporal information from cardiac musculature and valves by default - the principle regions of interest in all cardiac diseases. This enables the characterization of subtle regional aberrations in myocardial motion for downstream analyses, that traditional manually extracted echocardiographic measures fail to capture. Finally, our system can track features of importance with no additional human supervision in the form of outlines or labels. This enables rapid deployment of our methods to a diverse array of echocardiography-based problems, in an unbiased, and structure agnostic fashion.

The poor predictive value of contemporary clinical risk scores is well documented, and many of the component parameters are consequences rather than true predictors of RV failure. Most risk scores were either developed without internal cross-validation, or uniformly falter when applied to new datasets.^17,20,21,32^ More recently, some have attempted to use Bayesian methods on larger clinical registries to predict post-operative RV failure.^33^ Critically, pervasive issues with missing data resulted in severe class imbalances (2.7% RV failure, vs 97.3% normal patients) leading to gross overestimations of predictive power when relying on performance metrics insensitive to changes in class distribution.^34^ Further strengthening our work is the use of the latest standardized definitions of post-operative RV failure.^21,32^ We chose to dichotomize our primary outcome of RV failure to include only ‘severe or greater’ grades of RV failure. This was based on the significantly higher risk of mortality in these patients compared to those with moderate and lower grades of disease.^35^ We repeated the analysis using our AI pipeline trained on data with ‘moderate or greater’ grades of RV failure as the primary outcome. The performance remained similarly predictive, (AUC 0.855 (95% CI 0.812-0.899; n = 290 patients); Supplementary Fig. 1).

One of the advantages of analyzing ECHOs rather than using clinical surrogate parameters, is that it allows for a direct assessment of the heart. The literature surrounding the predictive value of manually calculated metrics of cardiac function remains inconclusive.^36,37^ In our study we identify that our radiomics AI echocardiography approach far outperforms manually calculated metrics of myocardial function in predicting RV failure. This further supports our rationale for moving away from segmentation or outlining based approaches for echocardiography AI, as automated calculations of the same metrics are unlikely to be predictive of our outcome of interest.

While our 10-fold cross validation demonstrated robust and stable performance of our AI system, our work in its present form is limited by the lack of a true external validation set. Our current efforts are directed at securing a large multi-center clinical and echocardiographic dataset for patients receiving LVADs to generalize our methods. Despite our relatively smaller sample size, making full use of the density of information in each video enables robust predictive performance. Prospective evaluations of predictive performance and integration with healthcare workflows are essential to understand the limitations of our technology in clinical practice.

We envision that by integrating AI systems within pre-operative clinical workflows, randomizing patients at high risk of developing post-op RV failure to various early right ventricular rescue strategies will finally be feasible. Our AI echocardiography tools may further serve as a clinical decision support system for instituting effective RV rescue treatments in this patient population. Our methods are, to our knowledge, the first to describe radiomics profiling for echocardiography analyses. This approach may find use in the early detection of heart failure, disease phenotyping, and a multitude of cardiac clinical decision support applications where treatment is guided by qualitative echocardiography assessments.

## Data Availability

Codebase for the AI platform will be made available along with all scripts used for statistical analysis upon peer-review. Patient level echocardiography data is not publicly available.

## Acknowledgements

This work is supported by a Stanford Artificial Intelligence in Medicine seed grant. Some of the computing for this project was performed on the Sherlock cluster. We would like to thank Stanford University and the Stanford Research Computing Center for providing computational resources and support that contributed to these research results.

## Supplementary Appendix

**Supplementary table 1:** INTERMACS definitions of RV failure. Updated RV failure definitions and index admission criteria for grading RV failure severity.

| <u>Right Ventricular Failure definitions:</u><br>Right Atrial Pressure > 16mm Hg.<br>**AND**<br>Hepatic (total bilirubin > 2.0 mg/dl) or renal dysfunction (creatinine > 2.0 mg/dl) |  |
| --- | --- |
| Severity Grade | Index Admission INTERMACS Criteria |
| **Mild** | Meets <u>both</u> criteria for RV Failure plus:<br>Post-implant inotropes, inhaled nitric oxide or intravenous vasodilators not continued beyond post-op day 7 following LVAD implant.<br>**AND**<br>No inotropes continued beyond post-op day 7 following LVAD implant |
| **Moderate** | Meets <u>both</u> criteria for RV Failure plus:<br>Post-implant inotropes, inhaled nitric oxide or intravenous vasodilators continued beyond post-op day 7 and up to post-op day 14 following LVAD implant |
| **Severe** | Meets <u>both</u> criteria for RV Failure plus:<br>Central venous pressure or right atrial pressure greater than 16mm Hg.<br>**AND**<br>Prolonged post-implant inotropes, inhaled nitric oxide or intravenous vasodilators continued beyond post-op day 14 following LVAD implant |
| **Severe Acute** | Meets <u>both</u> criteria for RV Failure plus:<br>Central venous pressure or right atrial pressure greater than 16 mmHg<br>**AND**<br>Need for right ventricular assist device at any time following LVAD implant<br>**OR**<br>Death during the LVAD implants hospitalization with RHF as the primary cause. |

**Supplementary table 2:** Scoring parameters for CRITT and Penn Score. For each criterion listed, a score of ‘1’ is assigned and the CRITT score is calculated as the sum of all components. For example, a patient with RAP > 15, tachycardia, and severe tricuspid regurgitation would have a CRITT score of 3. The Penn score is calculated by similarly assigning a score of ‘1’ for each criterion listed, the calculation however is (18 × CI) + (18 × RVSWI) + (17 × Creatinine) + (16 × Previous cardiac surgery) + (16 × Severe RV dysfunction) + (13 × SBp).

| Scoring System | Pre-operative parameters |
| --- | --- |
| **CRITT Score** | Right atrial pressure (RAP) > 15<br>Severe RV dysfunction (Qualitative ECHO report)<br>Severe Tricuspid Regurgitation<br>Tachycardia (HR > 100)<br>Ventilator requirement |
| **Penn Score** | Cardiac Index (CI) ≤ 2.2<br>RV stroke work index (RVSWI) ≤ 0.25<br>Creatinine ≥ 1.9 mg/dl<br>History of previous cardiac surgery<br>Severe RV dysfunction (Qualitative ECHO report)<br>Systolic blood pressure (SBp) ≤ 96 mm Hg |

**Supplementary Fig. 1:**
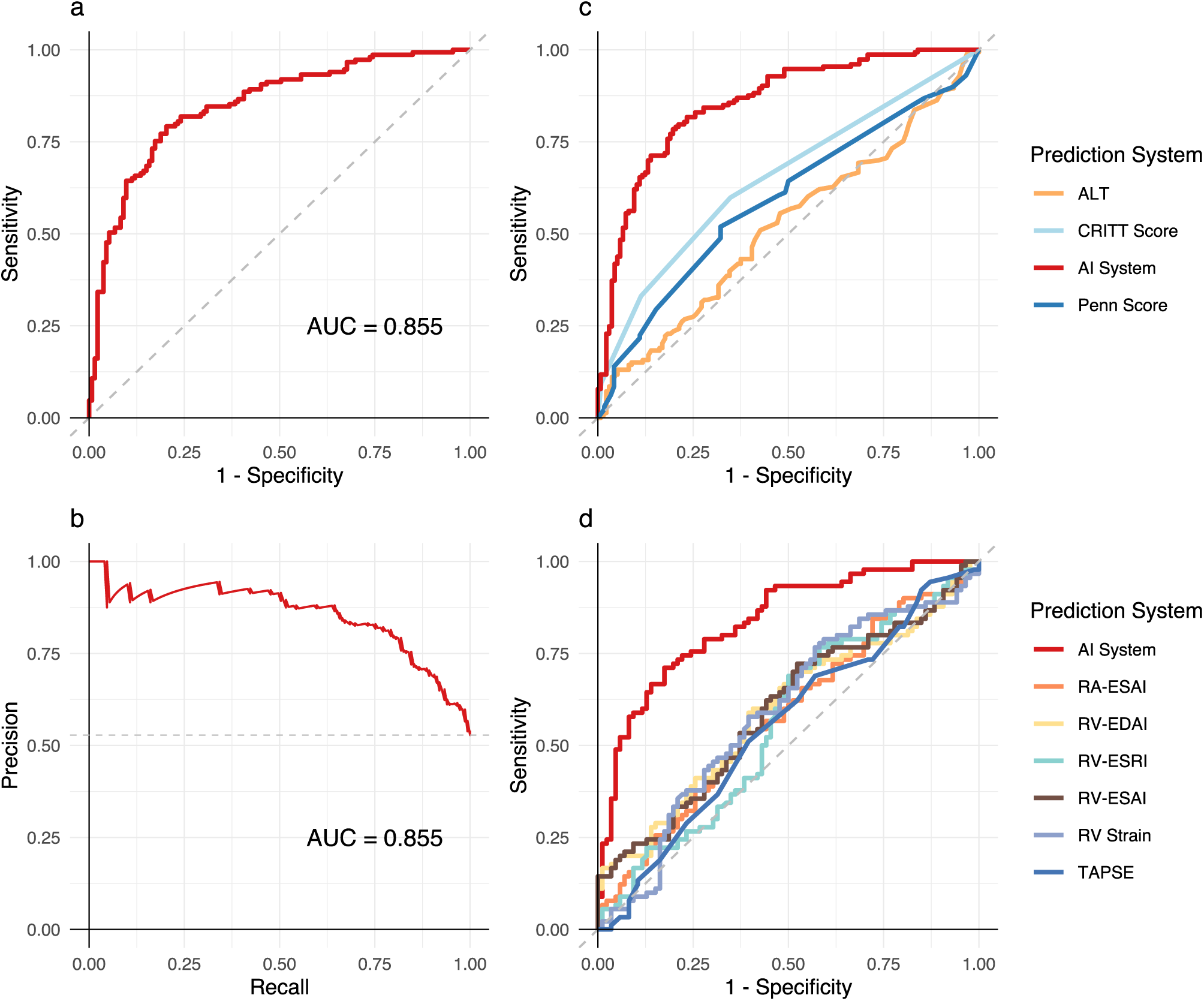
Performance of the AI echocardiography system, clinical risk scores, and manually calculated right heart functional metrics for RV failure outcome defined with a grade of ‘moderate or greater’. **a**. Receiver operating characteristic curve for the AI system; and **b**. Precision-Recall curve for the AI system (n = 290). The area under the ROC curve is 0.845 (CI = 0.803-0.893), this performance is similar to the AI system trained to predict ‘severe or greater’ grades of RV failure (p = 0.72) **c**. The ROC curve for the AI system compared to an independently calculated set of echo metrics for right heart function (n = 176) and **d**. the ROC curves of contemporary state of the art clinical risk scores (n = 229).

